# Lack of genetic evidence for NLRP3-inflammasome involvement in Parkinson’s disease pathogenesis

**DOI:** 10.1101/2023.09.20.23295790

**Authors:** Konstantin Senkevich, Lang Liu, Chelsea X. Alvarado, Hampton L. Leonard, Mike A. Nalls, Global Parkinson’s Genetics Program (GP2), Ziv Gan-Or

## Abstract

Activation of the NLRP3-inflammasome has been implicated in Parkinson’s disease based on *in vitro* and *in vivo* studies. Clinical trials targeting the NLRP3-inflammasome in Parkinson’s disease are ongoing. However, the evidence supporting NLRP3’s involvement in Parkinson’s disease from human genetics data is limited. In this study, we conducted analyses of common and rare variants in NLRP3-inflammasome related genes in Parkinson’s disease cohorts. We performed pathway-specific analyses using polygenic risk scores and studied potential causal associations using Mendelian randomization with the NLRP3 components and the cytokines IL-1β and IL-18. Our findings showed no associations of common or rare variants, nor of the pathway polygenic risk score with Parkinson’s disease. Mendelian randomization suggests that altering the expression of the NLRP3-inflammasome, IL-1β or IL-18, does not affect Parkinson’s disease risk or progression. Therefore, our results do not support a role for the NLRP3-inflammasome in Parkinson’s disease pathogenesis or as a target for drug development.

## Introduction

In recent years, activation of the nucleotide-binding oligomerization domain-, leucine-rich repeat and pyrin domain-containing 3 (NLRP3) inflammasome has been implicated in Parkinson’s disease by numerous functional studies using different models ^1^. Inflammasomes are protein complexes which serve as signaling platforms for activation of immune response. The NLRP3 inflammasome comprises three main components: NLRP3 (encoded by the *NLRP3* gene), apoptosis-associated speck-like protein containing a caspase activating recruitment domain (encoded by *PYCARD*) and caspase-1 (*CASP1*). NLRP3 is expressed in microglia and when activated, it leads to secretion of the cytokines IL-1β and IL-18, which leads to neuroinflammatory response and pyroptosis ^2^.

The evidence for the involvement of the NLRP3 inflammasome in Parkinson’s disease is mainly derived from *in vitro* and *in vivo* cell and animal models, by interacting with α-synuclein, mitochondria and other mechanisms. For example, early research suggested that in human monocytes, α-synuclein may directly trigger the NLRP3 inflammasome ^3^. Similar results have been reported in other cell and animal models ^4,5^. Other studies in cell and animal models have suggested that the NLRP3 inflammasome may be involved in toxin-mediated Parkinson’s disease and that there could be an interplay between mitochondria and the NLRP3 inflammasome in Parkinson’s disease pathogenesis ^6,7^. In humans, one study reported that a genetic variant in *NLRP3* may affect its expression and the risk of Parkinson’s disease ^8^. Several studies in cells and postmortem brain tissues from Parkinson’s disease patients and controls reported alterations in the NLRP3 inflammasome in Parkinson’s disease ^8–10^. However, there are no thorough human genetic studies of the NLRP3 inflammasome in Parkinson’s disease, although such studies can help with inferring causality. Nevertheless, there is a suggestion that the NLRP3 inflammasome may be a good target for therapeutic development in Parkinson’s disease, and several compounds targeting the NLRP3 inflammasome are in different stages of development ^11^. Considering that clinical trial success rates increase significantly when supported by genetic evidence ^12^, it becomes crucial to conduct thorough genetic analysis of the proposed target.

In this study, we aimed to examine whether human genetics data supports NLRP3 involvement in Parkinson’s disease and development of therapeutics targeting NLRP3 for Parkinson’s disease. We analyzed common and rare variants in the NLRP3 inflammasome components in large Parkinson’s disease cohorts, and further performed pathway specific analyses of polygenic risk scores (PRS) and Mendelian randomization (MR) analyses. Our results do not support an important role for the NLRP3 inflammasome in Parkinson’s disease nor its being a good target for therapeutic development in sporadic Parkinson’s disease.

## Results

### No association between NLRP3 inflammasome genes and Parkinson’s disease

We examined common variants from the largest available Parkinson’s disease risk GWAS (N cases/proxy-cases= 49,053; N controls= 1,411,006) ^13^. We did not observe any associations between Parkinson’s disease and variants in genes composing the NLRP3 complex (*NLRP3, PYCARD* and *CASP1*) and the genes encoding the cytokines released by its activation, *IL-1*β and *IL-18* in neither GWAS on participants from European ancestry nor the diverse non-European Global Parkinson’s Genetics Program (GP2) cohorts (Fig. 1; Supplementary Figure 1). While the *PYCARD* gene is located near one of the GWAS loci (rs11150601) within *SETD1A*, *PYCARD* is just outside of the linkage disequilibrium (LD) block, i.e. there are no variants within or in regulatory regions of *PYCARD* that are in LD (r2<0.2) with the variants that surpassed the GWAS level of significance. We then performed PRS analyses for the three NLRP3 inflammasome genes from 14,828 Parkinson’s disease cases and 13,283 controls across 7 cohorts (detailed in Supplementary Table 1). Overall, the PRS explains a very small portion of the variance in Parkinson’s disease (1.39E-06-0.001) and was not associated with Parkinson’s disease (Fig. 2, Supplementary Table 2).

**Figure 1.**
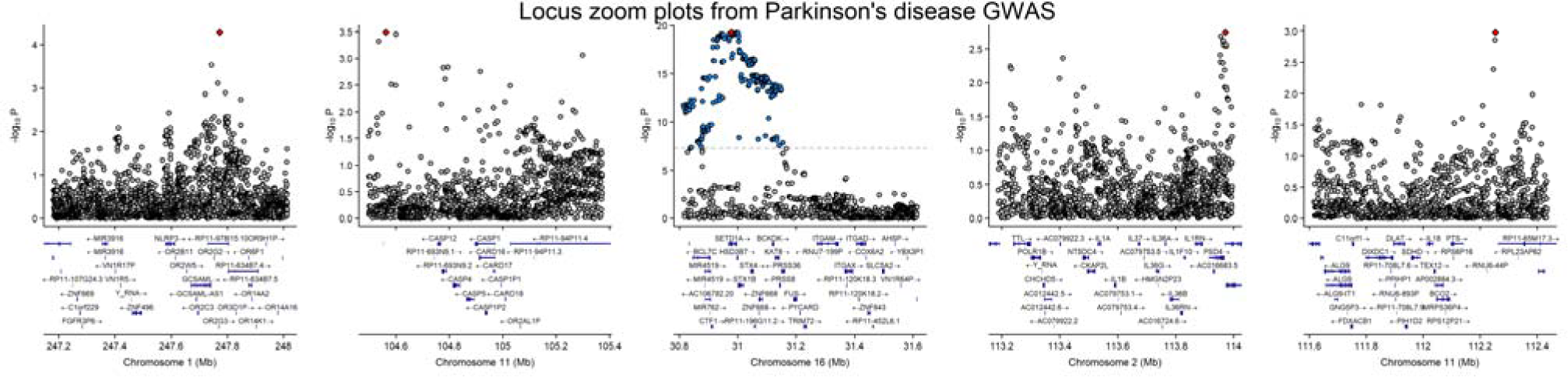
Locus zoom plot of *NLRP3*, *CASP1*, *PYCARD, IL-1*β and *IL-18* genes (+/-500 kb) in Parkinson’s disease GWAS. The SNP with the lowest P-value in the studied gene or locus is highlighted by red square. The punctuated line represents GWAS level of significance p < 5 × 10^−**8**^. SNPs shown in grey are below this significance level, while those in blue exceed it. X-axis: Chromosomal position (in Mb). Y-axis: Negative log10 of the P-values from GWAS.

**Figure 2.**
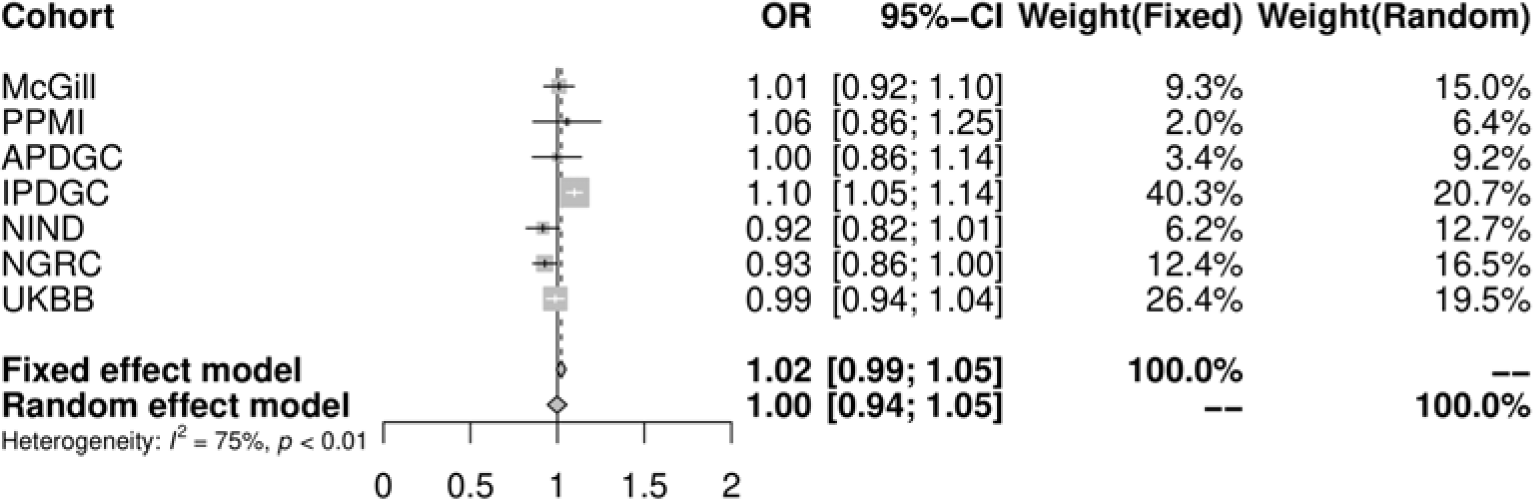
Pathway specific PRS analysis of NLRP3 inflammasome genes. OR-odds ratio; CI-confidence interval; PPMI-Parkinson’s Progression Markers Initiative; APDGC-Autopsy-Confirmed Parkinson Disease GWAS Consortium; IPDGC -International Parkinson Disease Genomics Consortium; NINDS-National Institute of Neurological Disorders and Stroke Repository Parkinson’s Disease Collection, NGRC-NeuroGenetics Research Consortium; UKBB-UK Biobank

We also analyzed rare variants in two independent cohorts, including 2,943 patients and 18,486 controls, followed by a meta-analysis (Supplementary Table 3). We did not observe any associations between any subsets of variants in any of the genes comprising the NLRP3 inflammasome and Parkinson’s disease (Supplementary Table 4). We then performed an analysis including all the variants in all three genes combined, and in this analysis too, we did not observe any associations between rare variants and Parkinson’s disease (Supplementary Table 4).

### Mendelian randomization does not support NRLP3 as a druggable target for Parkinson’s disease

Using summary-data-based Mendelian Randomization (SMR), we investigated whether the modulation of the NLRP3 inflammasome could be a target for therapy. Initially, we established that *NLRP3*, *CASP1*, *IL-1*β, *IL-18* are recognized within the database of druggable genes, highlighting their potential relevance for drug development ^14^. PYCARD, however, did not meet these criteria. We than performed SMR, where exposure were Quantitative trait loci (QTL) of NLRP3 genes from tissues that are relevant to Parkinson’s disease pathogenesis. In the present study, we used the Genotype-Tissue Expression (GTEx) project v8 release (All brain tissues, blood and liver), PsychENCODE, and BrainMeta/brain-eMeta ^15–17^. As an outcome for SMR, we used the most recent Parkinson’s disease risk ^13^, Parkinson’s disease age-at-onset ^18^ GWASs and largest publicly available Parkinson’s disease motor and cognitive progression GWASs ^19,20^. Our analysis did not reveal any potential causal associations between the QTL data tested in this study and Parkinson’s disease in tissues relevant for Parkinson’s disease after correction for multiple comparisons (Supplementary Table 5).

## Discussion

Our results, using large-scale human genetic, transcriptomic and methylomic data, do not support the NLRP3 inflammasome as important in Parkinson’s disease pathogenesis or as a good target for drug development. There were no associations of common or rare variants, nor of polygenic risk score for the NLRP3 inflammasome, with risk of Parkinson’s disease. When we considered the three NLRP3 genes as druggable targets, there was no evidence that altering their expression at the RNA level may have an effect on risk, onset or Parkinson’s disease progression.

While using MR to infer efficient druggability is not a definitive test, it can still provide valuable information. For example, a recent MR study was able to replicate the beneficial effects of tumor necrosis factor (TNF) inhibition in Crohn’s disease and ulcerative colitis, and its deleterious effect in multiple sclerosis ^21^. The same study also suggested that TNF inhibition might not be beneficial for Parkinson’s disease.

Understanding the role of a drug compound is essential when planning clinical trials. Studies that are not guided by genetic evidence are more likely to fail ^12^. Currently, several phase 1 clinical trials targeting neuroinflammation and particularly NLRP3-inflamassome are being conducted ^22^. The discordance between the hypothesis underlying these clinical trials targeting NLRP3 pathway in Parkinson’s disease and our findings suggests that efforts to target the NLRP3 inflammasome in Parkinson’s disease should be critically evaluated. It is important to select therapeutic strategies based on robust human genetic and biomarker evidence to reduce chances of trial failure. Perhaps targeting the NLRP3 inflammasome could work specifically in individuals in which this pathway is pathologically activated, but this approach is not being taken to the best of our knowledge. Subpopulations of patients with distinct genetic or environmental risk factors where the NLRP3 pathway plays a role in disease pathogenesis may exist. However, additional effort on defining subpopulation of Parkinson’s disease patients with neuroinflammation and particularly with induction of the NLRP3 pathway should be considered in clinical research targeting the NLRP3 inflammasome. Future studies exploring potential gene-environment interactions may further explain the role of the NLRP3 inflammasome in specific subsets of Parkinson’s disease patients.

Our study has several limitations that need to be acknowledged. The GWASs on Parkinson’s disease progression that were used could be underpowered. Further analysis using larger datasets should be performed when they become available to confirm our findings. Another important limitation of our study is the reliance solely on genetic and transcriptomic data to infer the role and druggability of the NLRP3 inflammasome in Parkinson’s disease. This approach does not account for post-translational modifications and the complex regulation at the protein level, which are critical for the functional activity of the NLRP3 inflammasome. Finally, the SMR analysis is dependent on the quality of the expression data used for exposure, and variations in quality across datasets might influence the results.

In conclusion, our analyses do not provide human genetic evidence for the involvement of the NLRP3 inflammasome in Parkinson’s disease, suggesting potentially limited druggability from genetic perspective.

## Online Methods

### Study populations

To examine whether common variants in the NLRP3 inflammasome components may be associated with Parkinson’s disease, we used summary statistics from the largest European Parkinson’s disease GWAS ^13^ and also analyzed genes of interest using the data from GP2 (release six) in several ancestry populations (detailed in Supplementary Table 6). Quality control analyses for both samples and variants have been previously described (https://github.com/GP2code/GenoTools). To study the association of common variants (minor allele frequency > 1%) with Parkinson’s disease in GP2 cohorts, we performed logistic regression, adjusting for age, sex, and the top five principal components in each ancestral population. For each of the cohorts, we created locus zoom plots using locuszoomr R library^23^ for the *NLRP3*, *CASP1*, *PYCARD, IL-1*β and *IL-18*, loci with +/-500kb around each gene. We then created pathway specific PRS for the NLRP3 inflammasome using available individual level data from cohorts of European ancestry (detailed in Supplementary Table 1).

In our Mendelian randomization analysis, we utilized the following summary statistics datasets: Parkinson’s disease risk GWAS ^13^, Parkinson’s disease age-at-onset GWAS with 17,415 cases ^18^, and Parkinson’s disease progression data from GWAS studies conducted by Iwaki et al. ^19^ and Tan et al. ^20^. The Parkinson’s disease progression traits in the study by Iwaki et al. ^19^ were measured using observational study meta-analysis of clinical scales data, we specifically used UPDRS Part III (N cases = 1,398), MMSE (N cases = 1,329), and MoCA (N cases = 1,000) scores. In the study by Tan et al. ^20^, Parkinson’s disease progression was assessed using scores for motor, cognitive, and composite progression in 3,364 Parkinson’s disease patients with an average follow-up of 4.2 years.

To analyze rare variants, we performed analysis in two cohorts with available whole-exome and whole-genome sequencing data with a total of 2,943 Parkinson’s disease patients and 18,486 controls (Supplementary Table 3). Whole-genome sequencing was available from the Accelerating Medicines Partnership – Parkinson Disease (AMP-PD) initiative cohorts (https://amp-pd.org/; detailed in the Acknowledgment). Whole-exome data was available from the UK biobank (UKBB) cohort which was accessed using Neurohub (https://www.mcgill.ca/hbhl/neurohub).

### Polygenic risk score pathway analysis

In order to examine the potential genetic association of the NLRP3 complex as a whole in Parkinson’s disease (as opposed to analysis of specific SNPs), we calculated pathway PRS using PRSet for the three genes encoding the components of the NLRP3 complex (*NLRP3, PYCARD* and *CASP1*) ^24^. In this analysis, we only included participants of European origin and removed first- or second-degree relatives. Sex discrepancy analysis was conducted to compare the recorded biological sex of individuals in the dataset with their genetically inferred sex, determined by rates of heterozygosity and homozygosity on the X chromosome. This analysis used the --check-sex function in PLINK 1.9, where males were expected to exhibit an X chromosome homozygosity estimate greater than 0.85, and females less than 0.25. This method enabled the identification and exclusion of samples with potential sex mismatches (where the reported sex at recruitment does not match the genetic sex), thereby enhancing the accuracy of subsequent genetic analyses. Only common SNPs with minor allele frequency > 0.01 and p-value < 0.05 were included in the analysis. We conducted LD clumping, removing variants with r2 > 0.1 and within a 250kb distance. We performed a permutation test with 10000 repetitions to generate an empirical p-value for our gene set of interest. We used age at onset for cases, age at enrollment for controls, sex, and the top 10 principal components as covariates.

### Whole-exome and whole-genome sequencing data analysis

To determine whether rare variants in the genes encoding the components of the NLRP3 inflammasome (*NLRP3, PYCARD* and *CASP1*), we extracted genetic data from whole-exome and whole-genome sequencing datasets. Our analysis included only participants of European ancestry, and we excluded any first or second-degree relatives from the study. For whole-genome sequencing data, we performed quality control as previously described ^25^. In brief, we included samples with a mean coverage of 25x and a rate of missing genotypes per sample less than 5%. For the UK Biobank’s whole-exome sequencing data, we used the Genome Analysis Toolkit (GATK, v3.8) to perform quality control. We applied the recommended filtration parameters for whole-exome sequencing data, which included a minimum depth of coverage of 10x and a minimum genotype quality (GQ) score of 20 ^26^. The human reference genome hg38 was used for alignment.

We analyzed the association of rare variants with minor allele frequency <0.01 using the optimized sequence kernel association test (SKAT-O) ^27^. The variants were grouped to different categories: all rare variants, all non-synonymous variants, loss-of-function variants (stop, frame-shift and canonical splice-site variants) and variants with a combined annotation dependent depletion (CADD) score ≥20 (representing 1% of the top deleterious variants). To meta-analyze the two cohorts we used the metaSKAT R package ^28^.

### Mendelian randomization

If modulation of the NLRP3 inflammasome is a target for therapy, then genetically driven differences in its expression, or that of the cytokines released following its activation, IL-1β and IL-18, should be causally linked to Parkinson’s disease risk or progression. To examine this possibility, we used SMR. SMR utilizes summary-level data to determine whether a causal relationship exists between an exposure and an outcome. In our specific case, we examined if differences in expression levels of the NLRP3 genes (using quantitative trait loci, QTL) are associated with risk, age-at onset and progression of Parkinson’s disease. As exposure, we used different QTL data from various studies and tissues including methylation, gene-expression and chromatin QTLs. All the QTLs we used were collected from the same resource, and we conducted analyses using SMR software developed by Yang Lab with standard settings (https://yanglab.westlake.edu.cn) ^29,30^. Specifically GTEx project v8 release (All brain tissues, blood and liver), PsychENCODE, and BrainMeta/brain-eMeta ^15–17^. As an outcome for SMR, we used the most recent Parkinson’s disease risk ^13^, Parkinson’s disease age-at-onset ^18^ GWASs and largest publicly available Parkinson’s disease progression GWASs ^19,20^. The Bonferroni-corrected significance threshold was set at p < 0.05/185=0.00027.

## Supporting information

GP2 Banner Author List

## Data availability

All code is available on our GitHub repository, which can be accessed at https://github.com/gan-orlab/NLRP3. The data used in the preparation of this article were obtained from the AMP PD Knowledge Platform (https://www.amp-pd.org) and the UKBB via Neurohub (https://www.mcgill.ca/hbhl/neurohub). The full GWAS summary statistics for the 23andMe inc., discovery data set will be made available through 23andMe to qualified researchers under an agreement with 23andMe that protects the privacy of the 23andMe participants. Please visit research.23andme.com/collaborate/ for more information and to apply to access the data. QTL data and SMR software are available on the Yang Lab website (https://yanglab.westlake.edu.cn).

## Acknowledgements

We would like to thank the participants in the different cohorts for contributing to this study. ZGO is supported by the Fonds de recherche du Québec - Santé (FRQS) Chercheurs-boursiers award, in collaboration with Parkinson Quebec, and is a William Dawson Scholar. The KS is supported by a clinical fellowship from the Parkinson Canada. Data used in the preparation of this article were obtained from the AMP PD Knowledge Platform. For up-to-date information on the study, visit https://www.amp-pd.org. AMP PD – a public-private partnership – is managed by the FNIH and funded by Celgene, GSK, the Michael J. Fox Foundation for Parkinson’s Research, the National Institute of Neurological Disorders and Stroke, Pfizer, Sanofi, and Verily. Genetic data used in preparation of this article were obtained from the Fox Investigation for New Discovery of Biomarkers (BioFIND), the Harvard Biomarker Study (HBS), the Parkinson’s Progression Markers Initiative (PPMI), the Parkinson’s Disease Biomarkers Program (PDBP), the International LBD Genomics Consortium (iLBDGC), and the STEADY-PD III Investigators. BioFIND is sponsored by The Michael J. Fox Foundation for Parkinson’s Research (MJFF) with support from the National Institute for Neurological Disorders and Stroke (NINDS). The BioFIND Investigators have not participated in reviewing the data analysis or content of the manuscript. For up-to-date information on the study, visit michaeljfox.org/news/biofind. The HBS is a collaboration of HBS investigators [full list of HBS investigators found at https://www.bwhparkinsoncenter.org/biobank/ and funded through philanthropy and NIH and Non-NIH funding sources. The HBS Investigators have not participated in reviewing the data analysis or content of the manuscript. PPMI – a public-private partnership – is funded by the Michael J. Fox Foundation for Parkinson’s Research and funding partners, including [list the full names of all of the PPMI funding partners found at www.ppmi-info.org/fundingpartners]. The PPMI Investigators have not participated in reviewing the data analysis or content of the manuscript. For up-to-date information on the study, visit www.ppmi-info.org. PDBP consortium is supported by the NINDS at the National Institutes of Health. A full list of PDBP investigators can be found at https://pdbp.ninds.nih.gov/policy. The PDBP investigators have not participated in reviewing the data analysis or content of the manuscript. Genome Sequencing in Lewy Body Dementia and Neurologically Healthy Controls: A Resource for the Research Community.” was generated by the iLBDGC, under the co-directorship by Dr. Bryan J. Traynor and Dr. Sonja W. Scholz from the Intramural Research Program of the U.S. National Institutes of Health. The iLBDGC Investigators have not participated in reviewing the data analysis or content of the manuscript. For a complete list of contributors, please see: bioRxiv 2020.07.06.185066; doi: https://doi.org/10.1101/2020.07.06.185066. STEADY[PD III is a 36[month, Phase 3, parallel group, placebo[controlled study of the efficacy of isradipine 10 mg daily in 336 participants with early Parkinson’s Disease that was funded by the NINDS and supported by The Michael J Fox Foundation for Parkinson’s Research and the Parkinson’s Study Group. The STEADY-PD III Investigators have not participated in reviewing the data analysis or content of the manuscript. The full list of STEADY PD III investigators can be found at: https://clinicaltrials.gov/ct2/show/NCT02168842. We would also like to thank the research participants and employees of 23andMe, inc. for making this work possible. The full GWAS summary statistics for the 23andMe discovery data set will be made available through 23andMe to qualified researchers under an agreement with 23andMe that protects the privacy of the 23andMe participants. Please visit research.23andme.com/collaborate/ for more information and to apply to access the data. This research used the NeuroHub infrastructure and was undertaken thanks in part to funding from the Canada First Research Excellence Fund, awarded through the Healthy Brains, Healthy Lives initiative at McGill University, Calcul Québec and Compute Canada. This research has been conducted using the UK Biobank Resource under Application Number 45551. The UKBB cohort was accessed using Neurohub (https://www.mcgill.ca/hbhl/neurohub). This research was supported in part by the Intramural Research Program of the NIH, National Institute on Aging (NIA), National Institutes of Health, Department of Health and Human Services; project number ZIAAG000535, as well as the National Institute of Neurological Disorders and Stroke. This work utilized the computational resources of the NIH HPC Biowulf cluster. (http://hpc.nih.gov). Data used in the preparation of this article were obtained from Global Parkinson’s Genetics Program (GP2). GP2 is funded by the Aligning Science Across Parkinson’s (ASAP) initiative and implemented by The Michael J. Fox Foundation for Parkinson’s Research (https://gp2.org). The members of the GP2 groups are listed in the supporting information. For a complete list of GP2 members see https://gp2.org.

## Supplementary material

Supplementary Table 1. Study population for pathway specific PRS analysis of NLRP3 inflammasome genes

Supplementary Table 2. Pathway specific polygenic risk score analysis on the of NLRP3 inflammasome in Parkinson’s disease

Supplementary Table 3. Study population for rare variant analysis

Supplementary Table 4. Burden analysis of rare variants in NLRP3 inflammasome related genes

Supplementary Table 5. Summary based Mendelian randomization studies between NLRP3 related genes expression loci and Parkinson’s disease risk and progression

Supplementary Table 6. GP2 study population for the role of common variants in NLRP3 inflammasome genes

